# Evolution during primary HIV infection does not require adaptive immune selection

**DOI:** 10.1101/2020.12.07.20245480

**Authors:** David A Swan, Morgane Rolland, Joshua Herbeck, Joshua T Schiffer, Daniel B Reeves

## Abstract

Modern HIV research depends crucially on both viral sequencing and population measurements. To directly link mechanistic biological processes and evolutionary dynamics during HIV infection, we developed multiple within-host phylodynamic (wi-phy) models of HIV primary infection for comparative validation against viral load and evolutionary dynamics data. The most parsimonious and accurate model required no positive selection, suggesting that the host adaptive immune system reduces viral load, but does not drive observed viral evolution. Rather, random genetic drift primarily dictates fitness changes. These results hold during early infection, and even during chronic infection when selection has been observed, viral fitness distributions are not largely different from *in vitro* distributions that emerge without adaptive immunity. These results highlight how phylogenetic inference must consider complex viral and immune-cell population dynamics to gain accurate mechanistic insights.

**One sentence summary:** Through the lens of a unified population and phylodynamic model, current data show the first wave of HIV mutations are not driven by selection by the adaptive immune system.

## Introduction

Longitudinal sequencing of HIV over time in an infected person provides invaluable insights into the pathogenesis of disease. Phylogenetic tools^1^ help illuminate evolutionary relationships between viral sequences observed at separate time points. Phylodynamic models leverage such relationships to further infer the virologic and immunologic forces governing observed viral evolution^2,3^. However, phylodynamic tools often do not capture the complex dynamics of HIV infection, which include massive exponential expansions and contractions of viral populations, mounting immune responses, target cell limitation, and existence of short- and long-lived cell populations. Mechanistic mathematical models of HIV explicitly include these complex, nonlinear interactions. To directly link mechanistic biological processes and evolutionary dynamics during HIV infection, we synthesized mathematical modeling and phylodynamics^4–10^ to design and validate a within-host phylodynamic (wi-phy) model of HIV primary infection.

## Results

### Viral dynamics and phylogenetics during primary HIV infection

We collected previously published data sets detailing three types of data relevant to early HIV infection: 1) non-linear viral dynamics during early HIV infection 2) viral evolution summary statistics including sequence divergence and diversity as well as tree-balance measures, and 3) HIV reservoir size and composition in terms of defective and intact sequences (see **Fig 1** & **Supplementary Table 1**). We then developed 9 fitting metrics to score model accuracy including viral kinetic measures (peak, set point, set point variability), evolutionary measures (divergence and diversity on day 20 and 40 after infection), and HIV reservoir sizes in intact and defective sequences (see **Methods** and all definitions and values in **Supplementary Table 2**).

**Figure 1.**
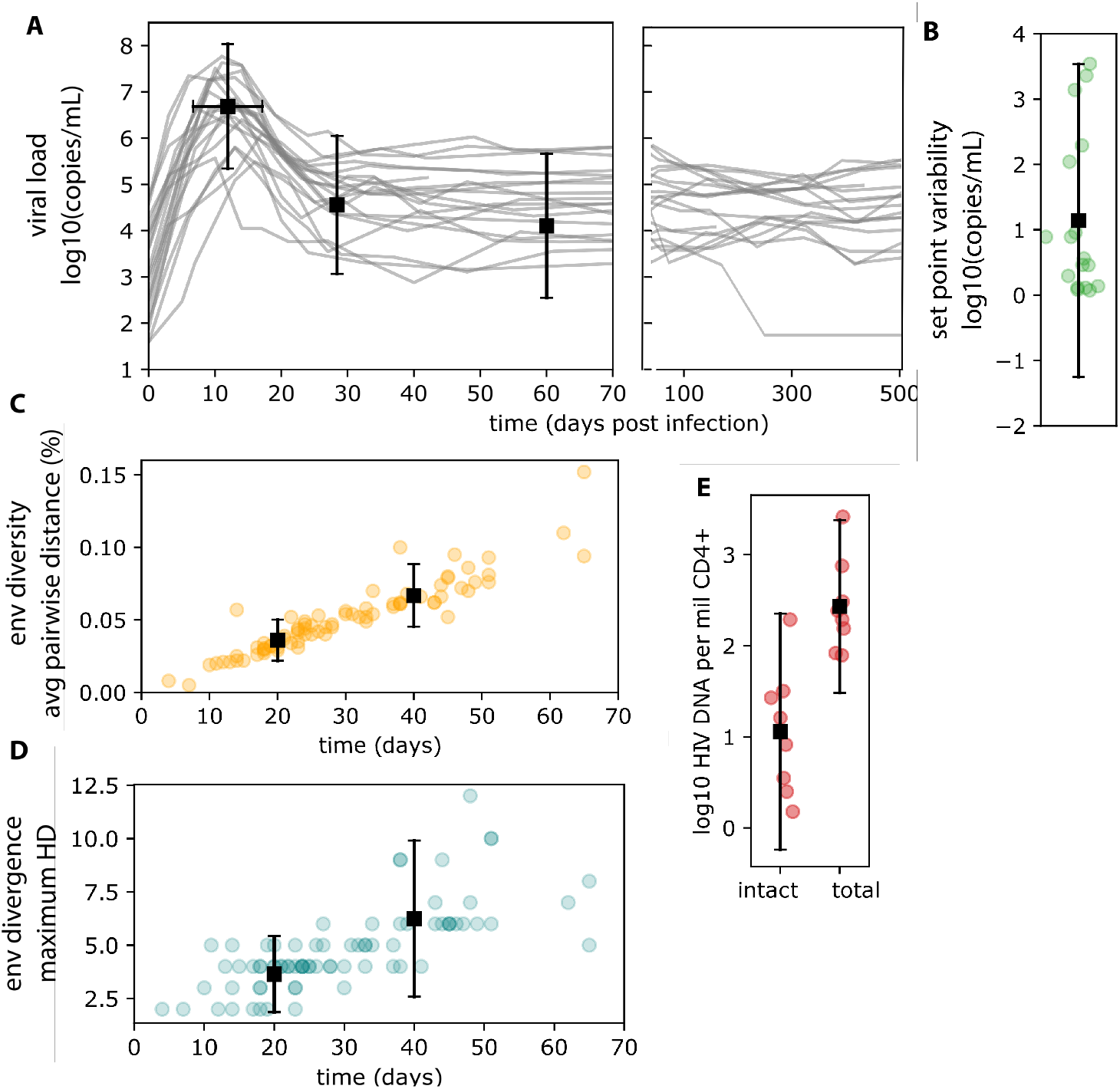
Phylodynamic fitting data. A) Viral RNA in blood from 39 individuals in Thailand and Uganda from the RV217 cohort. Box and whisker plots indicate mean and 95% CI for peak, nadir and setpoint viral load, as well as timing of peak (horizontal whiskers). B) Variation in setpoint viral load was quantified as the variance in viral load from participants with more than 10 longitudinal observations after day 50. C) HIV env nucleotide sequence diversity and D) divergence from 102 Clade-B HIV-infected individuals. E) Total and intact HIV reservoir measurements from 8 individuals in a UCSF cohort treated within 100 days of estimated infection. References and additional details are presented in **Supplementary Table 1**.

Because no individual has sufficiently granular data for all 3 types, we opted to fit to population data across types. This process also results in a balance toward higher variance of parameter estimates rather than a more accurate, but more biased parameter estimate as might arise from fitting to single individuals.

### Within-host phylodynamic model

The population dynamical model (**Fig 2A**) is a variant of the canonical viral dynamics model^11–13^ including virus, active and latently infected cells, and host adaptive immune response. Each viral variant *V*(*g*) is identified by its genotype (integer *g*). While the model is discrete and stochastic, the governing rules can be approximately expressed using ordinary differential equations (see **Methods**). Upon cell infection, the mutation rules (**Fig 2B**) are layered onto the model such that sequences may incur no mutations, point mutations, or mutations resulting in defective sequences (large deletions^14^ or hypermutations caused by a host anti-viral factors such as APOBEC^15^). We directly impute several important HIV-specific parameters including viral burst size (*π*=1000 virions per cell^16^) and clearance rate (*γ*=23 per day^11^), viral cytopathic infected cell death rate (*δ*_*A*_=1 per day^11^), HIV mutation rate (*μ*=3×10^−5^ per generation per base^17^), and the probability of latency (τ=10^−4^) as well as associated latent cell proliferation and death rates^18^.

**Figure 2.**
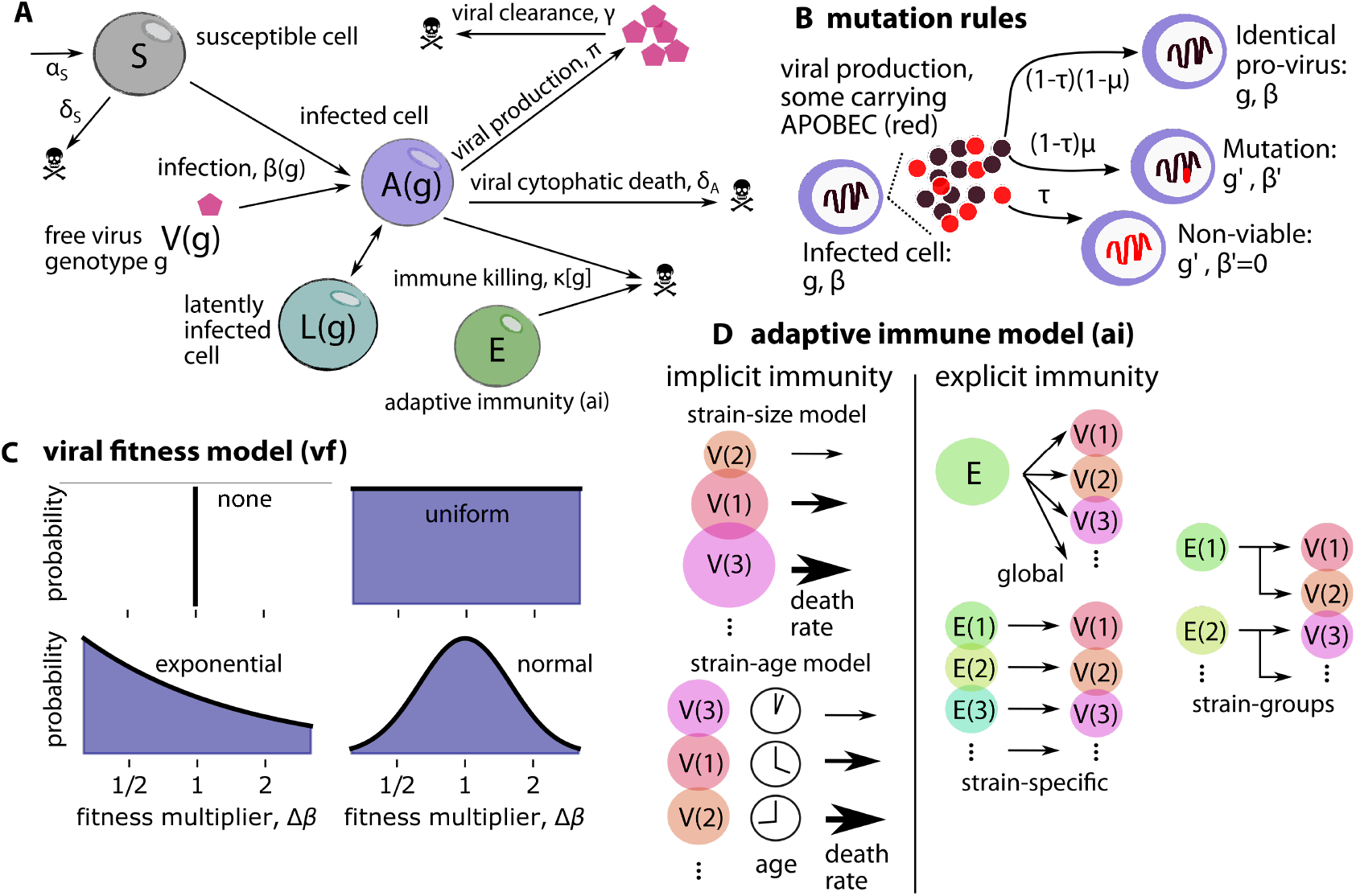
Schematic of mechanistic within-host phylodynamic (wi-phy) model. A) Population-dynamical model schematic. Susceptible cells are infected with a viral variant carrying a certain genotype V(g), which generates a new infected cell that can be active or latently infected. Actively infected cells produce virus and engender a host immune response; latent cells persist for years and probabilistically reactivate. B) Mutation rules govern the changes in viral fitness and Hamming distance to founder (HD) upon viral replication. Cell infection generally results in mutations (probability μ), though by chance sequences can remain identical (probability 1 − μ). Point mutations lead to new fitness (probability 1 − τ). We also allow for large deletions or hypermutations which nullify viral fitness (probability τ). C) A new sequence’s viral fitness is calculated by multiplying its parental fitness by a value drawn from the ‘viral fitness (vf) model’ distribution – tested models included none (all new viruses have the same fitness), and uniform, normal, and exponential distributions. D) The mechanistic form of the adaptive immune system was varied in 6 models with 3 main categories: 1) none, meaning no adaptive immunity at all 2) implicit, meaning that no adaptive immune compartment is modeled and death rate implicitly contains the response: 2a) variants are killed more if they have many associated cells (size) or 2b) have persisted for a long time (age), and 3) explicit, meaning that an additional state variable E(g) is used to model adaptive immune cells: 3a) global, all variants are killed by a single adaptive immune compartment, 3b) specific, each variant is killed only by its cognate adaptive immune compartment, and 3c) groups, collections of related variants are killed by an adaptive immune compartment.

### Simplifications to achieve computational tractability

Unlike many previous models that explicitly track viral variants as nucleotide sequences, we identify each variant as a unique genotype (an integer *g*) and track the complete transmission record as well as several key descriptive attributes for each genotype: fitness, Hamming distance to the founder sequence (HD), and age. As a result, we do not link viral genotype and phenotype, and fitness changes are not ascribed to mutation of a certain genomic locus. However, nucleotide sequences can be reconstructed from the transmission record to enable alignment and phylogenetic inference on sub-sampled data (see **Fig 6** and **Methods**). Another substantial enhancement was achieved by tracking population size but not attributes and transmission records for defective variants. This choice was valuable because hypermutants and/or large deletions are typically removed before analysis of experimental data, but modeling the number of defective sequences was crucial to accurately populate the latent reservoir, which is well known to be predominantly defective^19^. Together, these decisions allowed us to simulate large population sizes with good temporal resolution and properly incorporate rare variants (10 mL of blood and ∼10^9^ virions, Δt=0.01 day). The code is freely available (https://www.github.com/FredHutch/WiPhy_HIV).

### Mathematical model selection

In order to determine the mechanisms required to accurately match experimental data, we performed an extensive model selection procedure. For viral fitness (“vf”, **Fig 2C**), we tested 4 models: all new sequences have the same fitness (vf-identical), all new sequences have a randomly assigned fitness (vf-random), and two models where sequence fitness was inherited, either based upon an exponentially-(vf-exp) or normally-distributed (vf-normal) change Δ*β* from its parent sequence. For adaptive immunity (“ai”, **Fig 2D**), we tested 6 models: one with no adaptive immunity; two where immune pressure was implicit, either based upon the size of a certain sequence population (ai-size) or the length of time that a certain sequence existed (ai-age); and three where immune pressure was explicit – meaning a compartment of the model *E*(*g*) was added – either based upon a scenario where adaptive immune cells can kill any HIV sequence (ai-global), or adaptive immune cells have a specific cognate genotype which they can only kill (ai-specific), or adaptive immune cells can kill a range of genotypes (ai-groups). In the third category, a typical dynamical model for adaptive immune cells was included that allows adaptive cells to wax and wane based on the number of infected cells, with eventual saturation occurring if too many cells are generated (see **Methods** for equations). **Supplementary Fig 1A** tabulates all models and number of free parameters. Total RSS from the optimal parameter set is shown in **Supplementary Fig 1B**. For each model (4×6=24 total) we tested 50 parameter sets per free parameter (range: 2-8) and performed at least 10 stochastic replicates, amounting to ∼72,000 simulations of 100-day infections.

### An optimal model with non-specific immunity and inherited viral fitness

Ultimately, we defined the best model by its balanced fit to all 9 summary statistics rather than total error (all metrics RSS<2, **Supplementary Fig 1C**): the global adaptive immune model with exponential inherited fitness. In this model new variants are equally susceptible to adaptive immunity, which grows and equilibrates during the first few weeks of infection. That this version of the wi-phy model most accurately fit to data meant that: 1) child sequences must inherit part of parental fitness so that lineages can persist while also more often than not be worse off than parents so that strain replacement occurs; and 2) strain-specific adaptive immune pressure was not necessary to capture the major features of within-host HIV phylodynamics in early infection. Neither a model where viral strains implicitly lose fitness over time nor models with explicit strain-specific immunity matched the data as well. Rather than immune mediated selection sweeps, our model favors fluctuating viral fitness as the primary mechanistic driver of observed HIV evolution during primary infection. In model simulations, most new sequences were less fit than their parents: new point mutations were only beneficial 11% of the time and the average variant was less than half (0.47) as fit as its parent. Additionally, any fitness gains were conditional on the sequence being intact, which occurs in the minority (∼5%) of *all* mutations.

Three stochastic replicate simulations of the best model with the best parameter set is compared to all data metrics in **Fig 3**: viral load peak, nadir and set point **A**, set point variability **B**, sequence divergence **C**, sequence diversity **D**, and intact and defective latent reservoir size **E**. All metrics are captured successfully. Our fitting protocol used sequential metrics, fitting change from peak to nadir and setpoint rather than absolute value because these metrics are correlated naturally (which explains weaker fit to absolute nadir and setpoint).

**Figure 3.**
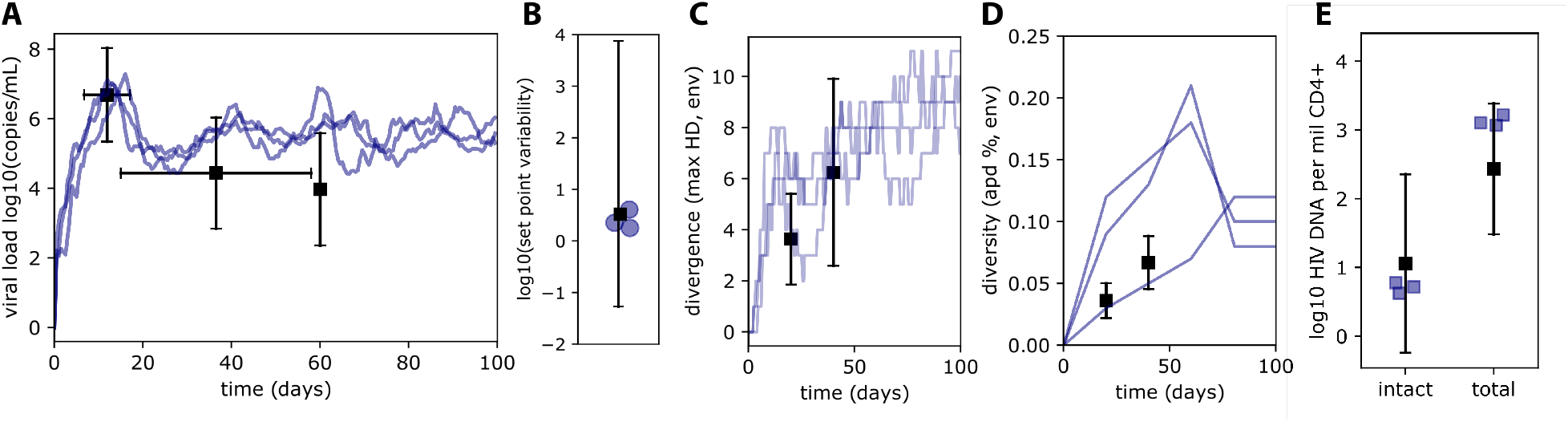
Stochastic phylodynamics predicted by the best-fit wi-phy model. 3 replicate simulations from the best-fit model (exponential fitness, global immune model) accurately recapitulate HIV viral population dynamics and phylodynamics: viral loads (A), set-point viral load variability (B), sequence divergence (C), sequence diversity (D), and intact and total HIV reservoir size (E). Dot and whiskers represent mean and 95% CI from data (See **Fig 1** and **Supplementary Table 2**). Stochasticity across replicates is evident and sequence sampling (to identical depths as experimental data) also introduces substantial variability.

### Data from chronic infection support the secondary impact of adaptive immune pressure

Next, we sought to test the model prediction that HIV phylodynamics can be explained in the absence of specific adaptive immunity in another setting. Thus, we compared the optimal viral fitness distribution against *in vivo* entropy distributions from sequences sampled later during infection. We collected data from the LANL database (https://www.hiv.lanl.gov) that quantify across 2339 individuals the relative abundance of each base *b* at each position *ψ* in the ENV gene such that a perfectly even distribution at some position is written *p*_*ψ*_ (*b*) = [0.25, 0.25, 0.25, 0.25], whereas a perfectly uneven distribution at some position is written *p*_*ψ*_ (*b*) = [0, 1, 0, 0], which represents that a single base (e.g. *T*) is found at that position for all individuals in the database. We calculated Shannon’s entropy *𝒮*_*ψ*_ = − ∑_*b*_ *p*_*ψ*_ (*b*) log *p*_*ψ*_ (*b*) for each position, which is shown in **Fig 4A**.

**Figure 4.**
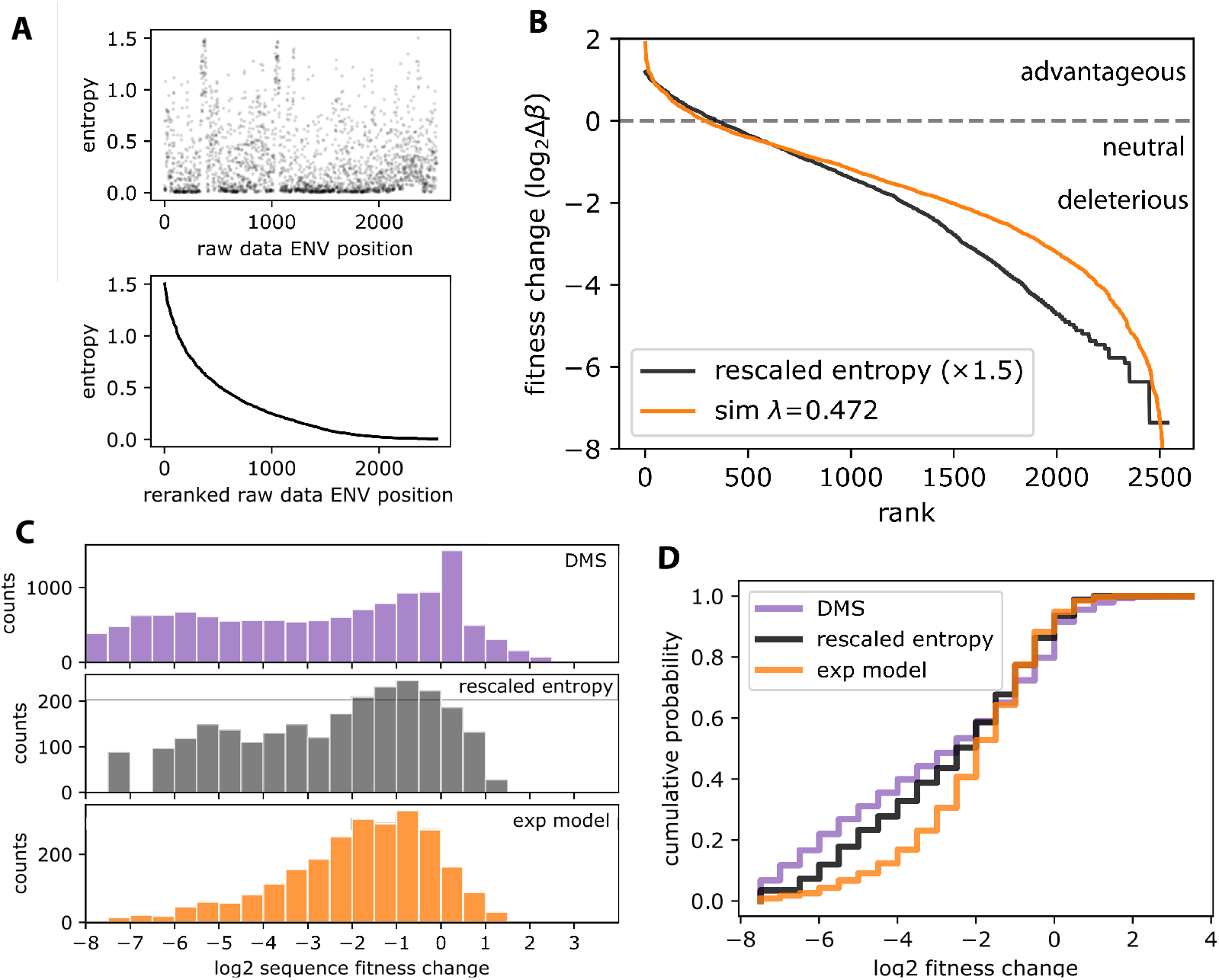
Fitness change distributions from chronic infection are similar to those of in vitro experiments performed without the influence of adaptive immune pressure. A) Available human data of ENV sequence position entropy (top panel), presented in rank order from most to least variable (bottom panel). B) Reranked entropy can be reimagined as the distribution of relative fitness changes – the most variable sites have low fitness costs when changed and the least variable sites have large fitness costs when changed. We use the model-derived fitness distribution (orange) to apply an absolute scale to the entropy distribution (rescaled ×1.5, black). We compare rescaled in vivo entropy to in vitro deep mutational scanning (DMS) data that quantified the relative fitness of all amino acid changes within a certain protein, here HIV ENV. We show histograms (C) and cumulative distribution function (D), to demonstrate that overlap between rescaled entropy and DMS data is reasonably similar. Notable differences occur at the peak of DMS, neutral (0) for DMS rather than slightly negative (−1) for entropy, and at the far tails of the distribution. The similarity of these distributions, both with 10-15% of mutations being advantageous, illustrates that during chronic infection, immune pressure does not select for particularly high fitness variants compared to those that exist intrinsically.

Next, because our model is agnostic to position-specific biology, we reranked entropy from most to least variable (**Fig 4A**, lower panel). We then use the distribution of entropy as a quantitative estimate of the relative effect of reducing fitness after a point mutation to positions in ENV. Positions with high entropy are flexible, and their fitness can increase or decrease given mutations. Alternatively, positions with low entropy lose fitness drastically if they are mutated. To put another way, we map the relative proportion of variants that exist in the database to the relative difference in fitness change after mutating one base vs another. Therefore, this distribution can be directly compared to the viral-fitness distribution in our wi-phy model. The relative difference in fitness change after mutating one base is precisely how we defined our model’s viral-fitness distributions. However, relative entropy has no scale, thus we identified the factor *x* that would scale entropy (assumed constant over position) such that *x𝒮* most closely resembled our best-fit viral-fitness distribution *p*(Δ*β*). We minimized the RSS between data and model distributions (**Fig 4B**) to find *x*∼1.5. Disagreement between model and data distributions occurred at larger fitness costs (Δ*β*<0.25). Fit was deemed less important in this range because variants experiencing large fitness costs, regardless of precise value, are subdominant and do not substantially influence data or simulations.

We next compared rescaled entropy with another data set: deep mutational scanning of HIV ENV^20,21^. These data in **Fig 4C** illustrate the fitness changes associated with *in vitro* perturbations of all amino acids in ENV. There is substantial agreement between this distribution and the rescaled entropy distribution of *in vivo* sequences (**Fig 4D**), indicating that *in vitro* fitness changes due to mutations are comparable to those observed during chronic infection *in vivo*. Both distributions contain a similar proportion (∼10-15%) of advantageous mutations. Therefore, large fitness enhancements *in vivo* appear less than or comparable to fitness enhancements *in vivo*, and could occur independently of T cell pressure. We cannot rule out the possibility that observed mutations are beneficial to the virus because they allow evasion of T cell pressure. Yet it again appears sufficient to describe phylodynamics without including that additional mechanism.

### Adaptive immune response has intense impact on viral load but limited impact on viral phylodynamics

To study the impact of the adaptive immune compartment further, we performed a global sensitivity analysis by simultaneously varying all model parameters and calculating the Spearman correlation coefficient between all parameters and all summary statistics (**Supplementary Fig 2A**). Importantly, of all parameters, the adaptive immune killing rate *κ* had the strongest impact on the drop to nadir and setpoint, illustrating the importance of the immune system to control viral dynamics especially after a high peak. Parameters regulating the maximal intensity of the immune response (saturation terms for both killing and recruitment, see **Methods** for equations) had relatively minimal impact on all metrics compared to other parameters. There was minimal correlation among adaptive immune parameters and phylodynamic measures (diversity and divergence at days 20 and 40). Instead, the average infectivity 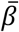 was the strongest determinant of phylodynamic metrics. Fitting score for phylodynamic measures show this pattern even more strongly (**Supplementary Fig 2B**). Together, these observations suggest that features of intrinsic viral fitness distributions have a primary effect on evolution and that adaptive immunity’s effect is secondary – modulating viral dynamics, which subsequently adjusts evolutionary dynamics. **Supplementary Fig 2C** shows predicted connections between population dynamic and phylodynamic measures that cannot be calculated from these data without a model (because they come from different individuals). In general, there was strong correlations among population dynamic measures and phylodynamic measures, but little correlation between these two broad categories. There was a notable lack of correlation between peak viral load and phylodynamics at or after day 20. While relatively weak, nadir and set-point viral load were correlated with phylodynamics, emphasizing the secondary impact of adaptive immune pressure on evolution through reduced viral load.

### Single variant viral dynamics during early HIV-1 infection

Using the best-fit model, we investigated individual variant viral dynamics by tracking the most abundant variants in a representative simulation (any variant entering the top 100 by abundance). First, we colored each variant by its genotype number (**Fig 5A**). During the first 3 weeks of infection there were only 1000 variants whereas by day 60, >300k productively infectious viral variants had been produced. Population sweeps visualized by the emergence of a new variant achieving top abundance and increasing diversity visualized by increasingly even abundances of concurrent sequences became evident at approximately day 40.

**Figure 5.**
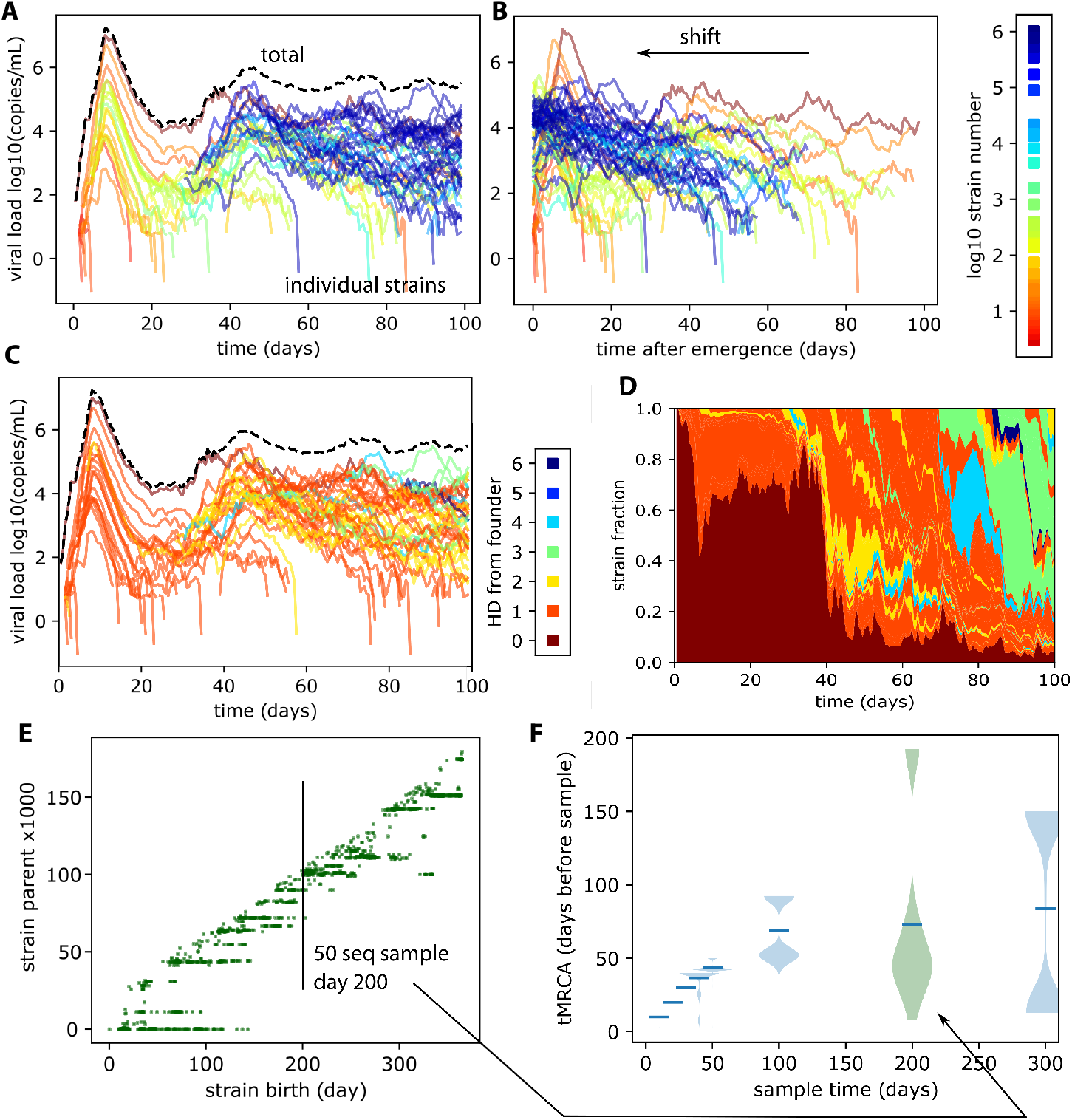
Visualizing evolutionary dynamics using the best-fit model. Using an example simulation of the best fit model (see **Fig 3** and **Supplementary Fig 2**), we plotted the total viral load as well as the viral loads from the top 100 most abundant variants. Our simulations are in 1 mL of blood which means that many more variants would be created in an infected individual than are observed in the sample. A) variants are colored by variant number, illustrating that population sweeps occur as red (strains 0-10) give way to yellow (strains 100-1000) etc. By day 40, almost 100,000 variants have been produced (i.e. blue variant). B) Shifting variant trajectories to start at the time they became dominant, we see that variants emerging later in infection have different kinetic profiles than those from early infection (compare red and blue). C) Coloring these same data by Hamming distance to founder sequence illustrates that many sequences persist for weeks with minor changes from the founder sequence. However, by day 40 after infection, sequential evolution has occurred, with variants emerging with 2-4 base pairs difference from the founder sequence. D) The proportion of dominant sequences illustrates the stark shift from founder predominance to a more balanced population after viral load nadir (same color scale as C). E) The parent of all variants can be visualized over time, showing a ladderlike evolutionary pattern. However, certain lineages continue for more than a hundred days, meaning that offspring are generated from a parental sequence that was created months prior. F) A random sample of 50 sequences taken at day 10, 20, 30, 40, 50, 100, 200, 300 and the times to most common ancestor (tMRCA) visualized as density plots. On day 200 the majority of sequences coalesce in the relatively recent temporal cluster (tMRCA∼10-40 days before sampling) while a minority arise from cocirculating lineages that persisted since early infection (tMRCA near 200). The bimodality of this distribution emerges between day 50-100, illustrating the waning influence of the founder sequence.

**Figure 6.**
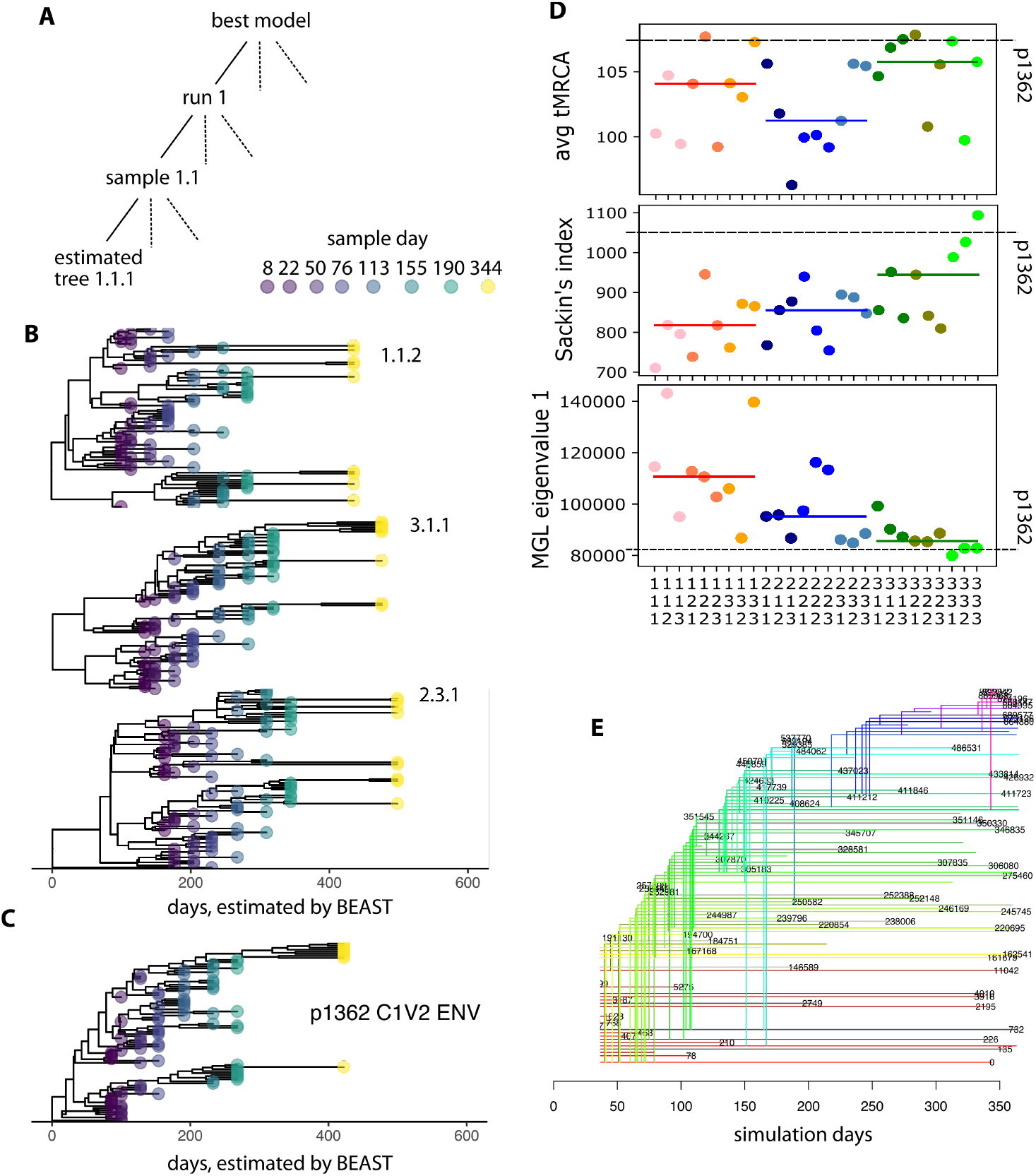
Comparative analysis of experimental and model tree estimation. A) The schematic for the tree inference testing process. We run the best model 3 times (i), for each we sample sequences with identical timing and sample size 3 times (j), and for each sequence sample, we perform 3 maximum clade credibility BEAST reconstruction of the phylogenetic trees (k). Therefore, we have 27 trees which we enumerate as i.j.k. B) 3 example estimated trees. Using a constant population size model leads to substantial overestimate of the time of infection prior to first sample. Here the first sample was at day 8 of infection, but the root was inferred to be ∼100-200 days prior, instead of 8 days to the founder sequence. C) A tree from an individual infected with HIV (C1V2 region of ENV; p1362). The sampling scheme in B is based on this individual. D) Several phylogenetic summary statistics express features of the evolutionary tree beyond diversity/divergence. These include time to most recent common ancestor (tMRCA), Sackin’s index of tree balance (sum of all internal nodes), and the modified graph Laplacian (MGL) largest eigenvalue, a surrogate for synonymous/nonsynonymous ratio. These results indicate that model runs introduce the most variability (mean; red blue green bars: i) relative to sampling or inference, (each identical colored dot represents an inference: k, and variants of red blue green colors indicate samplings: j). These results show the data from p1362 are relatively similar, even though our tree model was not built on this individual. Importantly, MGL eig 1 also falls within range of 1 simulation. E) A “true” tree from the sampled sequences illustrates all true sequence coalescence events taken from the complete transmission record in the simulation.

Next, by realigning variants to their time of emergence (setting *t* = 0 when the variant entered the top 100, **Fig 5B**), we identified that there were 2 dominant kinetic profiles. The first were those variants from before and during peak viremia, which have a large spike of >10^5^ viral copies (red/yellow); the second were those mostly generated after day 60 (when global adaptive immunity was appreciable), which peak at ∼10^4^ viral copies and slowly decay (blue).

Importantly, population sweeps were not caused by strain-specific targeting by the immune system (as models that include this mechanism were tested but not as successful, see **Supplementary Fig 1**). Instead, they were caused by continual mutations such that the currently fittest variant was unlikely to continue propagating without mutation and furthermore that subsequent mutations to this variant were equivalently likely to produce fit children as other newly circulating variants (all drawn from the exponential fitness change distribution with a minority of mutations being advantageous). This distinction emphasizes that sweeps, or strain replacement, again are sufficiently encoded by intrinsic viral fitness rather than asymmetric removal by adaptive immune pressure.

Coloring the variants instead by their Hamming distance to the founder virus (**Fig 5C**), we show that a starlike phylogeny dominates for approximately the first 40 days – meaning that while many distinct variants have emerged, they are all only 1 or 2 mutations away from the founder virus, and that the founder virus is thus the common ancestor of all. A shift from a starlike phylogeny arrives as sequential mutations build up and certain variants come to predominate with 3 or 4 base pair mutations distance from the founder around by 50.

The dominance of the founder virus for the first 40 days is also evident by examining variant fraction (calculated as the ratio of each variant viral load to the total, **Fig 5D**). When the founder loses predominance, the other variants are similarly competitive and thus a balance of several competing variants becomes more apparent. The timing of these results agrees with independent data showing shifts from demographic to selective effects around day 50^22^.

### Highly granular simulated phylogenetic trees

While phylogenetic trees require inference tools to visualize assumed viral evolution between observed sample timepoints, we have access to the complete transmission record from these simulations and can therefore examine evolutionary relationships with high granularity. In **Fig 5E**, each dot indicates the parental variant of child variants born on a certain day. In this way, we visualize how persistent parental variants were over time. For example, the founder variant (*g*=0) was very prolific, giving birth to new variants for months. Several other notable lineages arose where a parental variant continues to produce offspring for weeks/months before undergoing a sweep such that a new variant becomes dominant and produces offspring.

Notably, the persistence of lineages far outlasts the persistence of any single infected cell, whose average lifespan as input from previous mathematical modeling is approximately 1 day. These results were not a feature of latency as they persisted when we tested a model with no latent compartment. Nor is it a function of immune escape, there was no mechanism in this model whereby variants become more apparent (or not) to the immune system over time. Rather, lineage persistence is a probabilistic behavior balancing viral production and deleterious mutation rate. Indeed, some variants can persist at subdominant levels (**Fig 5E**). Here, dots at the same y-axis with gaps on the x-axis indicate times between which that parental variant was not dominant to the point of guaranteed sampling. Results were again repeatable without the latent cell compartment, suggesting that reactivation from latency does not typically drive subdominant persistence.

In **Fig 5F** we illustrate the distribution of times to most common ancestor (tMRCA) for a subsample of sequences (N=50, day 200). We calculate the time to common ancestor for all 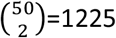 pairs, revealing a bimodal ancestry. Most sequences (∼90%, those that fall into the bars on the right side) had a relatively short tMRCA, coalescing to common ancestors approximately 10-40 days prior to the sampling date. This represents a time-localized quasi-species that is being generated actively by a dominant circulating variant. However, a minority swarm coalesced to an ancestor near acute infection which indicates some co-circulating lineages persisted since the time of transmission.

### Model validation with estimated phylogenetic trees

We next extended our model simulations out to 1 year of infection to test how typical experimental design and phylogenetic inference assumptions influence the interpretation of a phylogenetic data set. We subsampled from complete transmission records and built sequence alignments by starting with a reference sequence and performing random mutations based on Hamming distances (see **Methods**). Variability was quantified in each modeling step (simulation, sampling, and tree estimation). Three complete simulations with the best model were performed. Three samples were obtained from each simulation. Three repeated tree estimates were performed on each sampled data set (schematic in **Fig 6A**). This results in 27 phylogenetic trees (1.1.1 -> 3.3.3), from which 3 examples are illustrated in **Fig 6B**.

These trees immediately illustrate potential misclassification of phylogenetic inference in the absence of a population dynamic model. Here we have used the simplest assumption of constant population size in BEAST^23^. Because we know in reality viral load has peaked early in infection, the tree inference substantially overestimates the distance between the root and the founder sequence. That is, the purple samples that were observed at day 8 of infection are placed in the maximum clade credibility tree at 100 days since the most recent common ancestor. While such artifacts might be overcome with more complicated population dynamic models in BEAST, another challenge arises from building bifurcating rather than polytomic trees. Polytomic trees allow for a single variant to produce many different offspring variants, exactly what occurs in our simulation as an infected cell produces many new mutated offspring that have a single common ancestor – thus additional internal nodes inferred by a bifurcating tree may be artifacts.

**Fig 6C** is an estimated tree made from experimental data with identical sampling scheme. **Fig 6D** illustrates 3 different phylogenetic summary statistics computed from each simulated tree and the experimental tree. Statistics are average tMRCA, Sackin’s index (a tree balance statistic calculated as the sum over the number of internal nodes between root and tip for all tips in the tree)^24^, and the dominant eigenvalue of the tree’s modified graph Laplacian (MGL) spectrum. MGL is a robust measurement of tree shape that quantifies deep/shallow branching events and importantly was shown to be a surrogate for synonymous to nonsynonymous (dN/dS) ratio, a metric often used to quantify selection^25^.

Across metrics, the variability introduced by sampling and tree estimation was substantial. However, only model runs has a significant effect on median values for these metrics (see median lines taken across sampling and tree estimation, different hues of orange/blue/green), particularly average tMRCA. The third run was most like the experimental tree in all statistics. These results suggest that phylogenetic inference tools may provide different conclusions given the same set of underlying viral sequences in blood, due to under sampling and stochasticity associated with the tree estimation approach. While impossible to access in real data, **Fig 6E** illustrates the true tree, using the accurately dated common ancestors of all sampled sequences available from the complete transmission record.

## Discussion

In this work, we developed the most data-driven model of within-host HIV primary infection phylodynamics to date. Modeling dynamics and evolution is computationally challenging, and was achieved through several key simplifications. First, to balance computation time and still allow rare discrete events we model 10 mL of plasma (∼2-3 orders of magnitude below that in an adult human). Next, we ignored nucleotide sequences until post facto analysis after sub-sampling. Instead we record the complete transmission record with each variant given a genotype number. This genotype specifies the variant’s fitness, age, and Hamming distance to the founder. Finally, while total population size is tracked, the transmission record only includes intact sequences (∼5% of all), which is realistic because many HIV phylodynamic studies discard hypermutated or sequences with large insertions or deletions before performing phylogenetic reconstruction^26^.

Through rigorous model selection against collected data sets, we found the most parsimonious model of HIV had two important characteristics that inform HIV pathogenesis. It was governed by 1) a distribution of fitness changes drawn from an exponential distribution such that most children are less fit than parental sequences and 2) an adaptive immune system that was equally potent against all variants.

The model viral fitness distribution was applied to rescale *in vivo* sequence entropy data and make it comparable to *in vitro* data on mutational fitness. In doing so, we found that the *in vivo* and *in vitro* distributions were not significantly different. Because there is no immune selective pressure *in vitro*, these results corroborated our view that much of HIV mutation is controlled at the viral rather than adaptive immune level.

Together these findings paint a picture of what is sufficient to describe the first year(s) of HIV infection: a viral quasi-species in which a fit variant can dominate or co-circulate with other dominant strains. However, the children of currently dominant variants are probabilistically likely to be less fit such that new variants emerge and take over. Such population sweeps are sufficiently modeled without any additional pressure from the immune system against specific variants.

In building simulated trees we also highlighted several challenges of tree estimation from real data in which depth and granularity of sampling is limited. Bearing these complexities in mind, we advocate for inclusion of population dynamic data whenever possible and continual enhancement of phylodynamic methods such as ours to disentangle these exquisitely coupled processes in practice.

There is strong evidence pointing to HIV-selection by adaptive immunity during chronic infection. These range from fixed mutations that can be linked to detectable CD8+ T cell response^27^, a dose-response relationship (in 1 individual) between immune pressure and escape rate^28^, an increase over time of escape mutations in HLA-matched hosts relative to HLA-mismatched hosts^29^, and the emergence of broadly neutralizing antibodies^30,31^. Our results do not invalidate these findings. Instead, in the context of the “red-queen phenomenon”^32^, it may be that sequential viral/host co-evolution is not particularly relevant for early pathogenesis.

Other implications from our results include that adaptive immunity may be broader than previously imagined and that its influence on evolution may be secondary, through modulating viral dynamics. The latter is in keeping with the fact that the timing of CD8 expansion correlates with reduction in viral load^33^ and cell depletion experiments in SIV infected macaques that indicate CD8+ T cells control viral load^34^ (for this point, we note other studies show inefficient infected cell killing by CD8+ T cells, suggesting a more nuanced interpretation^35^). Additionally, because the effector cell killing rate *κ* determined viral load setpoint, we suggest that the overall HLA-antigen match, which depends on the specific host *and* the specific virus determines disease severity. This agrees with the finding that certain host HLA genotypes are associated with delayed progression to AIDS^36^, but that the founder sequence is correlated to pathogenesis^37^. Importantly, while the intrinsic viral fitness distribution drives evolution in our model, adaptive immunity is still required to control viral levels thus affecting evolution in a secondary manner.

Our modeling has several limitations. The magnitude of our modeled adaptive immune response cannot be directly compared to existing data from the various studies because *E*(*g*) is not precisely representing any specific cell type (e.g. CD8+ T cells, anti-HIV antibodies, natural killer cells), and likely only captures the HIV-specific arm of the immune system. The fitness landscape of HIV fitness costs has been modeled in more detail previously^38^. Susceptible cells are also not clearly defined phenotypes. In ∼15% of cell infections viral progeny share genetic material from 2 parental sequences that infected the same cell^39,40^. We do not explicitly simulate such genetic recombination. While explicit modeling of recombination could be added as described previously^41,42^, mutational distance is small during early infection such that we do effectively allow for larger changes (potentially from recombinants) by allowing for >1 point mutation per cell-infection. We do not attempt to model compartmental anatomy, instead relying on past studies that show HIV dynamics are reasonably consistent across tissues^43–45^.

Finally, genotype and phenotype are not directly linked at present, meaning we cannot model sequence-specific phenomenon (nor compare to structural biology data or simulate ART-escape mutations). We cannot therefore model the important quantity of nonsynonymous to synonymous ratio (dN/dS). This ratio has been used to demonstrate selective pressure as dN/dS>1 suggests mutated nucleotides that meaningfully change proteins (nonsynonymous mutations) are more likely to fix. Still, for our conclusions, dN/dS>1 is not obvious in the first years of infection^46^ and additionally, the MGL summary statistic (a surrogate for dN/dS^47^) agreed between our model-derived phylogenetic trees and human trees sampled in the first year of infection.

By unifying viral population dynamics and phylodynamics in this model, we have shown that primary HIV infection is not driven by adaptive immune selection as strongly as previously thought. This has important ramifications for vaccine design attempting to target certain epitopes and speaks to evolutionary steering^48^ (of the virus into a less fit state) and broadly neutralizing antibodies to supplement the broadly killing adaptive immune system.

Future applications of the wi-phy model abound. From testing phylogenetic inference on simulated data, to estimating infection timing, to modeling therapies with broadly neutralizing antibodies and/or predicting phylogenetic signatures of the reservoir after curative interventions. As within-host viral genetic data are continuously collected, this type of model is crucial for their further interpretation.

## Data Availability

All data are available from previous studies and model code is freely available upon request.

## Acknowledgments

This work was funded by a WRF postdoctoral fellowship and an NIAID K25 to DBR. DBR is grateful to numerous colleagues for conversations including Florencia Tettamanti-Boshier, Trevor Bedford, Bethany Dearlove, Eric Lewitus, Pavitra Roychoudury, and particularly Paul Edlefsen and Jim Mullins who were instrumental in initial conceptualization of this model.

## Materials and Methods

### Mathematical description of the model

We implement our model stochastically, but we express the model as ordinary differential equations for readability (see **Fig 2**). It contains cells susceptible to HIV infection, *S* which are created with rate *α* and die with rate *δ*_*S*_. HIV infection begins with the introduction of a founder HIV sequence with genotype *g* as an intact actively infected cell 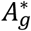 (superscript ∗ denotes intactness). This cell produces virions *V*_g_ which infect more cells with rate *β* _g_ *SV*_g_. When a new cell is infected, mutations occur with rate *μ*, or not with probability (1 − *μ*). We use the measured HIV mutation rate of 3.5 × 10^−5^ nucleotide substitutions per generation^17^. Given mutation, the proviral sequence is chosen to be intact (with probability τ) or defective (probability 1 − τ). Intact sequences are typically generated by point mutations. A small proportion of infected cells enter a latent state 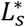 with the small probability *λ*_*S*_ where subscript *s* further subdivides latent classes to satisfy observed multi-phasic decay. Latent cells proliferate, die and reactivate to an active state with rates *α*_*S*_, *δ*_*S*_, *ξ*_*S*_ respectively. Thus, we have many possible infected cell states, which for brevity we express as *I*_g_.

The rules of the mechanistic model can be expressed as a growing set of differential equations (*∂*_*t*_ denotes time derivative). After each time step, new sequences are added by mutation such that {*g*} → {*g, g*′}.

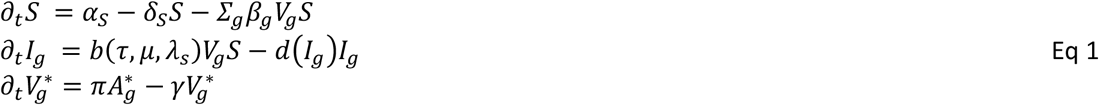

The generic creation and removal rates of each type of infected cell is governed by the term *b* and *d*, such that for example, the creation of active, mutated, intact infected cells is given by *b* = (1 − τ)*μ*(1 – *∑*_*S*_ *λ*_*S*_). The generic removal rate of infected cells depends on their state and genotype *d*(*I*_*g*_), and is varied to allow for the various adaptive immune models in the model selection process.

To track evolutionary distance to the founder sequence we modify the Hamming distances of mutated sequences by drawing a Poisson distributed number of base pair mutations with average 1,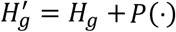. Defective sequences are typically generated through hypermutation or large insertions/deletions, which may be caused by a host anti-viral factor like APOBEC3G. We modify these Hamming distances by drawing a Poisson distributed number of base pair mutations 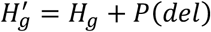, where 40 is the average number of base pair changes for hypermutations^17^.

### Viral fitness (vf) models

We created 4 models for the viral fitness (vf) of mutated intact sequences. The first is a trivial model where each viral strain has the same fitness. Thus *p*(Δ*β*) = *d*(Δ*β*) where *d* is the Dirac delta function equal to zero unless Δ*β* = 1. The second assumes no inheritance from parental strains such that fitness changes are uniformly distributed with a maximum value *p*(Δ*β*) = *U*[0, *β*_max_]. The third and fourth models assume heritability of viral fitness, either with an exponential distribution *p*(Δ*β*) = exp(−*k*Δ*β*) /*k* or a Gaussian distribution *p*(Δ*β*) = 𝒩 (1, *σ*_β_). However, in both cases, we also enforce the constraint that *β*_g_ ′ ∈ [0, *β*_max_].

### Adaptive immune (ai) models

The genotype dependent death rate of actively infected cells *d*(*A*_g_) is used to incorporate 6 models of the adaptive immune (ai) response. The first model has no adaptive immunity such that *d*(*A*_g_) = *δ*_*A*_*A*_g_.

The next 2 models have ‘implicit immunity’, meaning that there is no additional compartments to represent immune cells. In the strain-size model, 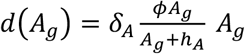. We interpret this as that the number of infected cells carrying that sequence effectively attracts immune cells and disproportionately kills it off with a multiplier *ϕ* on the natural death rate *δ*_*A*_ and a saturation constant *h*_*A*_. In the strain-age model, *d*(*A*_g_, *t*) = *δ*_*A*_ exp[*r*(*t* – *a*_g_)]*A*_g_. We interpret this as that the older sequences (larger time between birth date *a*_(_ and time after infection *t* have had more time to accrue adaptive immune pressure, and thus are eliminated more rapidly. This model enforces strain replacement with rate constant *r*.

The next 3 models contain explicit immunity. First, we add a state variable compartment representing effector cells *E*_g_(*t*). This part of the biology is the least understood and our mechanistic implementations may effectively capture several types of cells or molecules (CD8+ T cells, NK cells, antibodies). We then add the differential equation,

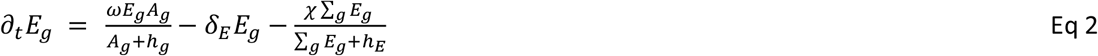

Depending on the model, effector cells have their own genotype *g* which matches a viral genotype. Then, cells grow based on the prevalence of their cognate antigenic genotype (term with nonlinear growth rate *ω* and saturation constant *h*_g_), die naturally with rate *δ*_E_, and have another death term such that the total adaptive immune response (sum over genotypes) is constrained in size (term with saturation constant *h*_g_).

In the global model, *d*(*A*_g_) = [*δ*_*A*_ + *κ* ∑_g_ *E*_g_]*A*_g_. We interpret this as there is a single adaptive immune compartment that can kill any strain.

In the strain-specific model, *d*(*A*_g_) = [*δ*_*A*_ + *κ*_g_*E_g_*]*A _g_*. We interpret this as for each viral strain there is an adaptive immune compartment that can kill only that strain. The killing ability of each strain specific adaptive immune compartment is *κ _E_*.

In the strain-group model, *d*(*A*_g_) = [*δ* _*A*_ + *K* ∑ _g∈i_ *E*_g_]*A*_g_. We interpret this as that there is some cross immunity so sequences with similar sequence numbers (within a group *i* ranging from 1 → *G*) can be killed by a single immune compartment with killing rate *K* which is assumed the same for all groups.

### Simulation implementation

The model is implemented in C++ and is freely available. We use a discrete stochastic τ-leap simulation scheme in 10 mL of plasma and a simulation time interval of Δt=0.01 days. The state variables ***X*** = {*S, A* _g_, *L* _g_, *V* _g_, *E*} represent the numbers of susceptible, active/latent infected cells and virions (for each genotype), and adaptive immunity if explicitly in the model. Thus, in each time interval, a Poisson distributed number of events of each mechanistic transition is chosen by the reaction propensities *p*_***X***_^49^ such that ***e*** = *Poiss*(*p*_***X***_Δ*t*). Then, the state variables are updated using the event transition matrix ***T*** as Δ***X*** = ***eT***. For example, in an interval, we might observe the creation of a new latently infected cell of a new genotype by viral infection. For this example, *T* = [*S* − 1, …, *L* _g_′ + 1, …, *V*_g_ – 1], meaning removal of a susceptible cell, removal of a virion of genotype *g*, and the creation of a latently infected cell with genotype *g* ′. In this same interval many other events could occur simultaneously.

### Tracking the transmission record

To capture evolution, each viral strain (or variant) is described by a genotype number (an integer *g*) which is specified by its age (*a*_g_ the date of its emergence in time since the start of infection), the number of base-pair mutations for this genotype relative to the founder virus (the Hamming distance *H*_g_), and a viral fitness (*β*_g_ – not necessarily unique). The number of each state variable associated to that strain is tracked so that we know the number of intact virions/infected cells/latent cells of each genotype at all times. The complete transmission record is tracked as the links between all genotype numbers to their parental genotype number. Importantly for computational tractability, defective strains are not included in the transmission record as they do not have offspring.

### Parameters from the literature

By using previous estimates, we constrained the parameters that must be estimated from 18 in the complete model to approximately 5. Experimentally derived parameters include the average number of virions produced during an infected cell’s lifetime (*π*)^50^, the clearance rate of free virus (*γ*)^16^, the proliferation and death rates of T cell subsets (*α*_c_, *δ*_c_)^51,52^, the probability of abortive infection (τ)^53^, and HIV mutation rates (*μ*)^17^. Other parameters have been estimated by mathematical modeling, including the death rate of infected cells (*δ*_*A*_)^54–56^, mean infectivity of the virus 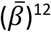, the killing rate of CD8+ T cells (*κ*)^57,58^, the rate of reactivation from latency (*ξ*)^59,60^, and the probability of HIV latency (*λ*_*S*_)^61^.

### Model fitting procedure

For each model we tested 50 values of each free parameter such that more complex models had more total simulations for optimization. The values were drawn from a grid search evenly spaced between a lower and upper bound for each parameter (often several orders of magnitude) based on previously determined HIV model rates. We then repeated 10 replicate simulations for each parameter set. The best replicate was used to score the parameter set using RSS against all metrics. That is, we scored each metric *M*_i_ as the value of the model *m*_i_ against the 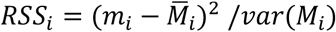 using the mean and variance of the metric. Then, we can calculate the total RSS as the sum *RSS* = ∑_i_ *RSS*_i_.

### Model selection

Because of the multiple (possibly correlated) metrics, it was unrealistic to assume that the sum of the individual metric scores would provide a proper likelihood. Thus, we ruled out the ability to score using typical model selection procedures using information criterion (e.g. AIC). Instead, we ranked all models by the best total score, and then investigated the top models to see which had the best balance against all metrics (see **Supplementary Fig 1**).

### Calculation of diversity and divergence

Using previous definitions^62^, we calculate the viral sequence diversity using the average pairwise distance of sampled variants. These calculations do not require a precise nucleotide sequence, but rather depend on our knowledge of the exact transmission record and the phylogenetic distance between sequences (known from their Hamming distances, respectively). To simulate sampling, we randomly select virions (recapitulating experimental sampling of viral RNA). To compare, we replicate sample sizes from all experimental studies used. To calculate the distance between 2 sequences, or the number of base pair differences, we compute the sum of their Hamming distances and subtract off the Hamming distance of their parental sequence Δ(*i, j*) = *H*_i_ + *H*_j_ – *H*_p(*i, j*)_. The maximum divergence is calculated as the max Δ(*i, j*) where *i* = 0 is the founder sequence. To calculate diversity, we calculate all pairs of Δ(*i, j*) for all *i* and *j* in a sample. We account for multiple sampling of the same variant *N*_*i*_ > 1, by the frequency of each sampled variant 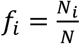, where *N* is the total sample size. Again, while we do not require the complete nucleotide sequence, we specify the number of base-pairs in the sequences *n*_*b*p_ (for env). Finally, the diversity is 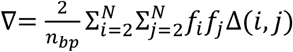.

### Calculating tMRCA

In **Fig 4** we illustrate a true tree and calculate the times to most recent common ancestor (tMRCA) for sampled variants. This is accomplished by examining all sequence pairs and determining their parental sequence. If the parent is identical, the procedure halts and the birth date of the parent is recorded as the time of most recent common ancestor. If the parent is different, we examine parents of the parental sequences and repeat. Eventually the common ancestor is found – sometimes leading back to the founder sequence.

### Entropy from patient data

We obtain filtered HIV1 Env alignment (type M w/o recombinants) from the LANL website and remove all but subtype B sequences resulting in 2339 sequences. Entropy is calculated with default options on the LANL website and gaps are removed to resolve the consensus sequence and its entropy values.

### Modeling nucleotide sequences

To integrate the model with phylogenetic inference tools, we took model simulations and added a nucleotide substitution model. This model begins with a benchmark HXB2 sequence, and applies mutations based on the Hamming Distance and parental lineage from the simulation output. We then apply an HKY nucleotide substitution model so that model output is amenable to Bayesian viral phylodynamics (BEAST)^23^. For more complicated post-facto sequence reconstruction, we can make mutations more likely to cause transversions vs translations by also specifying for each genotype the number of each nucleotide bases (*ACGT*).

### Sampling and integration with BEAST

At the given interval, cells are sampled from the pool of virus present until a given number of sequences are represented or until all active sequences are represented if the actual number is less. The sequence index of each sampled cell is output to the log file. The variants sampled and the number of cells picked for each variant during this process are used when calculating the diversity of that sample at the sample time. Diversity is calculated using pair-wise variant distances to the most recent common ancestor. That ancestor is the actual one from the variant lineage table and not one estimated via analytic tools as might be done with a real sample. Divergence is calculated by looking at the maximum hamming distance to the earliest common ancestor across all variant pairs. Thus, we do not automatically assume a star-like phylogeny. After the simulation has completed, the log file, the FASTA file for the founder variant’s gene of interest and saved variant lineage table are used together to reconstruct the sequences present in each sample at each time point. This is possible because each mutation during simulation run time is assigned a set of one or more transitions (A->G, G->C, etc.) based on the A/C/G/T mix of the parent variant and the probabilities for each type of transition. (Note: this strategy could be expanded to include insertions and deletions, but not recombination). The earliest sample time for each sequence is used to construct a time-labeled FASTA file for the samples which can be used as input to phylogenetic analysis tools such as BEAST.

**Supplementary Table 1.** Data characteristics.

| Fitting metric | Data type | Population sample size | Individual time-points | Sequence sample size | Sequence type | Stage of infection | Reference |
| --- | --- | --- | --- | --- | --- | --- | --- |
| **Viral load** | Viral RNA, Clade A/E&C | 39 | 10 | >10 | NFL | Primary | Robb et al. NEJM 2016 |
| **Sequence divergence** | Viral RNA, Clade B | 102 | 1 | median 25, (range=10-203) | ENV | Primary/ Chronic | Keele et al. PNAS 2008 |
| **Sequence diversity** | Viral RNA, Clade B | 102 | 1 | median 25, (range=10-203) | ENV | Primary/ Chronic | Keele et al. PNAS 2008 |
| **Reservoir** | DNA | 8 | 1 | $\sim 10^6$ CD4+ cells | NFL | On ART | Bruner et al. Nat Med 2016 |
| **ML Tree** | Plasma RNA PID1312 | 1 | 8 | 30 μl Viral RNA | C1V2 ENV | Primary/ Chronic | Liu et al. JVI 2006 |

\*NFL, near full length; ML, maximum likelihood

**Supplementary Table 2.** Model fitting metrics. Each metric was calculated for each individual, and the population average and variance are presented.

| Metric | Definition | Value: Avg (Var) | Units |
| --- | --- | --- | --- |
| **Peak VL** | Maximal viral load within 30 days of estimated date of infection | 6.7 (0.77) | log10 copies/mL |
| **Peak-nadir difference** | Minimal viral load after peak and before day 40, calculated as change relative to peak | 2.38 (0.59) | log10 copies/mL |
| **Set point VL** | Average viral load during set point, defined as 10 days after nadir to final observation | 4.44 (0.88) | log10 copies/mL |
| **Set point variability** | Standard deviation of set point VL, using all observations during set-point | 0.14 (0.04) | log10 copies/mL |
| **Peak time** | Time of maximal viral load, relative to first positive | 12.24 (11.58) | days |
| **Nadir time** | Time of nadir, relative to first positive | 26.87 (77.68) | days |
| **Diversity** | Average pairwise distance between sequences | 0.172 (0.0013) |  |
| **Divergence** | Maximum Hamming distance (base pair differences) relative to observed founder sequence | 17.56 (52.97) |  |
| **Defective fraction** | The ratio of defective to all sequences | 0.95 |  |

**Supplementary Figure 1.**
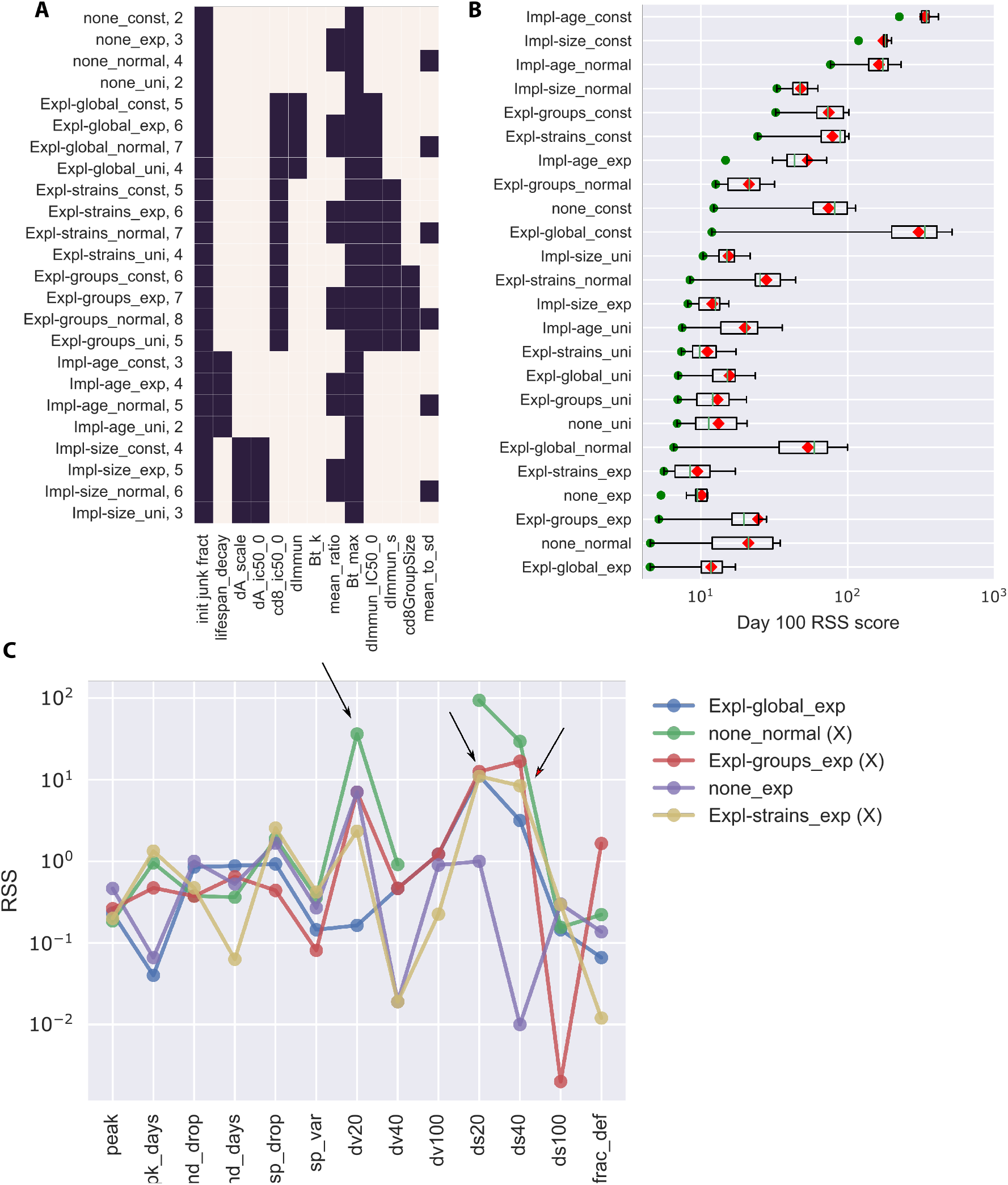
Model selection. We tested 24 models with 50 parameter sets per free parameter (see A) and 10 replicate simulations per parameter set. For each model, the best replicate simulation of the best parameter set was used to score the model by RSS. In (B), we illustrate the best run (green dot) as well as the range (boxplot) and mean (red diamond) from the best parameter set of each model. This illustrates that some models have more stochasticity across replicates than others. The models are ranked from best to worst by the best single replicate (lowest to highest RSS, bottom to top). To better study the top 5 models, we plotted the RSS against each metric, and eliminated models with RSS for any metric above 10. Note the log10 scale here, models with RSS<1 are excellent fits.

**Supplementary Figure 2.**
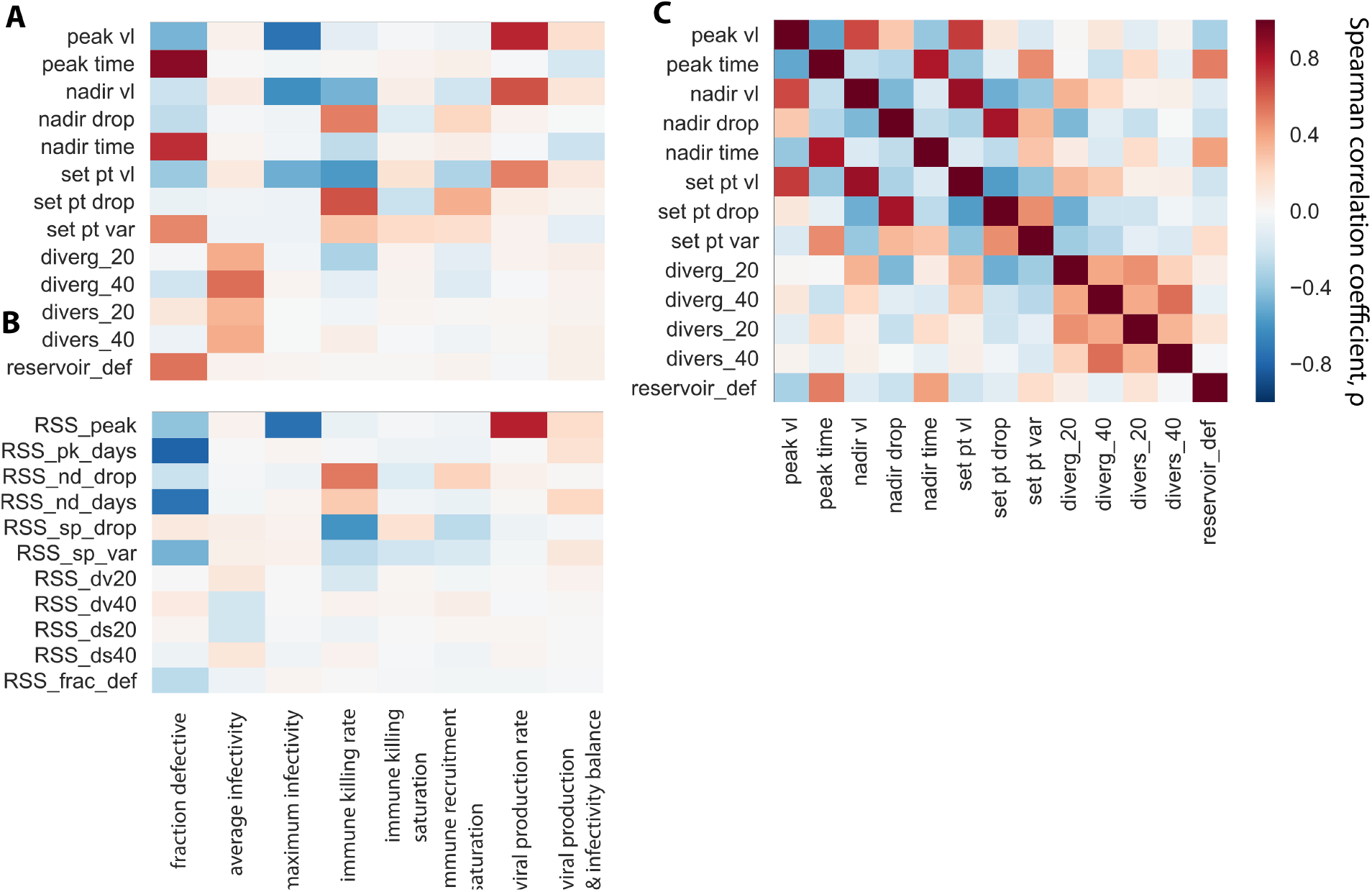
Global sensitivity analysis reveals phylodynamic correlates during HIV primary infection. Using the selected model, we performed a global sensitivity analysis of 300 parameter sets, and compared each free parameter in this model (8 total) against the values of metrics (A) and the scores of against the metrics (B). Results are discussed in the main text. Additionally, we note that the fraction of sequences that are defective τ was strongly correlated with several metrics. Intuitively, raising τ made sequences in the reservoir more likely to be defective, and it is the only model parameter that adjusts this metric. However, τ also effectively adjusts infectivity, such that more defective sequences reduced peak viral loads and delayed peak and nadir. τ also increased the variability of set point viral load because it reduces the number of intact lineages, making total viral load fluctuate more with stochastic gains or losses of a single lineage. Maximum infectivity was required in the model to avoid new lineages emerging and effectively creating another peak viral load. This parameter counterintuitively correlated with population dynamic values: raising it lowered all of peak, nadir, and set point viral load. This may be partially due to an artifact of simulation where runs with unrealistically high viral loads (>10^9^ copies/mL) are terminated for computational reasons. Viral production rate conversely affected the same metrics. Intuitively, increasing the number of virions created by a single infected cell in its lifetime raises peak, nadir, and set point viral load. C) Using the selected model, we performed a global sensitivity analysis of 300 parameter sets, and correlated population dynamic and phylogenetic summary statistics. These serve as predicted phylodynamic correlates during HIV primary infection.

